# A novel artificial lung organoid for simulating a patient derived adenocarcinoma of lung for personalized oncology

**DOI:** 10.1101/2021.04.20.21255803

**Authors:** Sally Esmail, Wayne R Danter

## Abstract

Optimizing patient care based on precision oncology will inevitably become the standard of care. If we accept the principle that every persons’ cancer is different then the most effective therapies will have to be designed for the individual patient and for their tumors genetic profile. Access to tumor mutational profiling is now widely available but continues to be limited by cost and actionable information. For example, novel combinations of approved drugs are rarely considered. These considerations lead us to hypothesize that artificially induced Lung Adenocarcinoma (LUAD) derived lung organoids could provide a novel, alternate approach for LUAD disease modeling and large-scale targeted drug screening.

In this project, we used data from a commercially available tumor mutation profile to generate and then validate the artificially induced LUAD-derived lung organoid simulations (aiLUNG-LUAD) to model LUAD and identify several drug combinations that effectively reverse the tumors’ genotypic and phenotypic features when compared with placebo. These results complement previous LUAD-derived lung organoids research and provide a novel and widely applicable cancer drug-screening approach for precision/individualized oncology.

## INTRODUCTION

Lung cancer is the leading cause of cancer-related death globally (Safiri et al., 2020). In 2020, Lung cancer accounted for 1.80 million deaths worldwide (Piñeros et al., 2021). Lung cancer can broadly be classified into Non-Small Cell Lung Cancer (NSCLC; ∼85%) and Small Cell lung cancer (SCLC; ∼15%). The main subtypes of NSCLC are adenocarcinoma, squamous cell carcinoma, and large cell carcinoma. Lung adenocarcinoma (LUAD), the most common type of NSCLC, is a highly heterogeneous disease, whose treatment remains challenging. Nevertheless, as precision oncology continues to advance, molecular gene profiling is widely employed to determine the best treatment options for individual patients (Politi and Herbst, 2015).

### LUAD disease modeling, Molecular profiling, and targeted therapies

Molecular profiling of adenocarcinoma has led to the discovery of EGFR mutations and ALK-rearrangements. This discovery and others have led to the first clinical use of cancer targeted therapies, namely tyrosine kinase inhibitors (TKIs) (Politi and Herbst, 2015). Additionally, the remarkable sensitivity to TKIs observed in some lung cancer patients suggested that profiling molecular subsets of lung cancer could guide further characterization of the lung cancer genome and led to the incorporation of molecular testing into routine clinical practice enabling precision oncology and targeted therapies research(Brown and Elenitoba-Johnson, 2020). Nevertheless, the development of individualized targeted therapies for LUAD has been limited by the lack of reliable and cost-effective *in vitro* models to enable high-throughput drug screening. Recently, the *in vitro* three-dimensional cancer-derived organoid models have been developed from different cancers including LUAD (Kim et al., 2019; Shi et al., 2020; Weisberg et al., 2020). A patient derived LUAD in vitro lung organoid has also been successfully developed and then used for evaluating tumor specific single drug therapies (Li et al., 2020c). However, creating individualized patient-derived cancer organoid is often hindered by the high cost of their laboratory establishment and validation for large-scale therapeutic discovery studies. Thus, we hypothesize that using artificially induced LUAD-derived lung organoids will provide a novel alternative disease modeling approach for LUAD disease modeling and large-scale targeted drug screening (Li et al., 2020c).

In this study, we have generated and validated artificially induced LUAD-derived lung organoid simulations (aiLUNG-LUAD) to model LUAD and enable biomarker discovery and high-throughput targeted-drug screening. This study complements previous LUAD-derived lung organoids research (Kim et al., 2019; Li et al., 2020c; Shi et al., 2020; Weisberg et al., 2020) and provide a novel cancer drug-screening approach for precision and individualized oncology.

## METHODS

The DeepNEU stem cell simulation ML platform is a hybrid learning system based on elements of fully connected recurrent neural networks (RNN), cognitive maps (CM), support vector machines (SVM) and evolutionary systems (GA). Our DeepNEU platform is built around a continuously expanding database of concepts and relationships that are derived from genomic/proteomic and pathways data gathered from the peer reviewed literature. The information in the database is stored in the form of an N by N square matrix with matrix values ranging between -1 for a maximally negative relationship to +1 for a maximally positive relationship. In this system 0 is used to represent an arbitrary reference point between maximal values. The basic fuzzy logic (FL) representation is easily converted to a Neutrosophic logic (NL) (Smarandache, 2010) by simply representing relationships that remains unknown as “Indeterminant” with the letter “I”. An initial state input vector is used to represent the state of the system at time zero. Iterative sparse vector matrix multiplication cycles continue until a new system wide, steady, or final state is achieved. The initial state vector can be modified to represent a wide range of gene-based disorders by turning off or on any combination of gene, protein, or phenotypic concepts. Our machine learning platform uses an unsupervised learning approach. In this case, unsupervised learning means there are no data profiles associated with specific outcomes and the system learns relationships between individual concepts rather than a specific number of outcomes. In our approach, unsupervised learning would be closer to clustering and regression than classification. The detailed methodology for simulation development and validation plus the description of the evolving DeepNEU database used in these experiments has been described previously in detail (Esmail and Danter, 2021a).

These proof of concept, personalized cancer treatment, DeepNEU based experiments, employed a five-step methodology. These steps are (1) development of wild type lung simulations (aiLUNG), (2) development of the adenocarcinoma of lung simulations (aiLUNG-LUAD), (3) validation of generated simulations, (4) single and multiple drug combination screening, and (5) statistical analysis of the generated data.

The literature validated DeepNEU platform is an artificial stem cell and organoid hybrid deep learning system with components from fully connected recurrent artificial neural networks (RNN), cognitive maps (CM), genetic algorithms (GA) and support vector machines (SVM). The methodology for simulation development and literature validation plus the description of the immediate predecessors (i.e. v6.1, v6.2) of DeepNEU v6.3 used in these experiments has been described in detail elsewhere in (Esmail and Danter, 2021a, b).

### (1) Developing the wild-type lung simulations (aiLUNG)

The primary goal of this project was to advance our DeepNEU based research into aiPSC and organoid simulations by evaluating the potential of the platform to simulate a patient derived cancer mutational profile and then deploy that simulation for identifying potential cancer drugs for repurposing. To that end, we first created computer simulations (aiPSC) of human induced pluripotent stem cells (iPSC) and then use the aiPSC to derive lung (aiLUNG) organoids. The recipe used to generate the aiLUNG organoids is summarized in Table 1 below. These aiLUNG wild type organoids were designed to simulate a wide range of lung cell types including Alveolar Type I (ATI) and Alveolar Type II (ATII) precursors, ATI and ATII cells, ATI and ATII Saccular cells plus epithelial Ciliated, Club, Goblet cells, pulmonary neuroendocrine cells (PNEC) as well as Lung Basal Stem Cells (BSC). Since it has been recognized that LUAD may originate from more than one cell lineage, this diversity of lung cell types is important to ensure that the potential cell or cells of origin of the tumor are represented in the organoid (Cheung and Nguyen, 2015; Ferone et al., 2020; Rowbotham and Kim, 2014). All experiments were carried out in triplicate using different initial conditions.

**Table 1:**
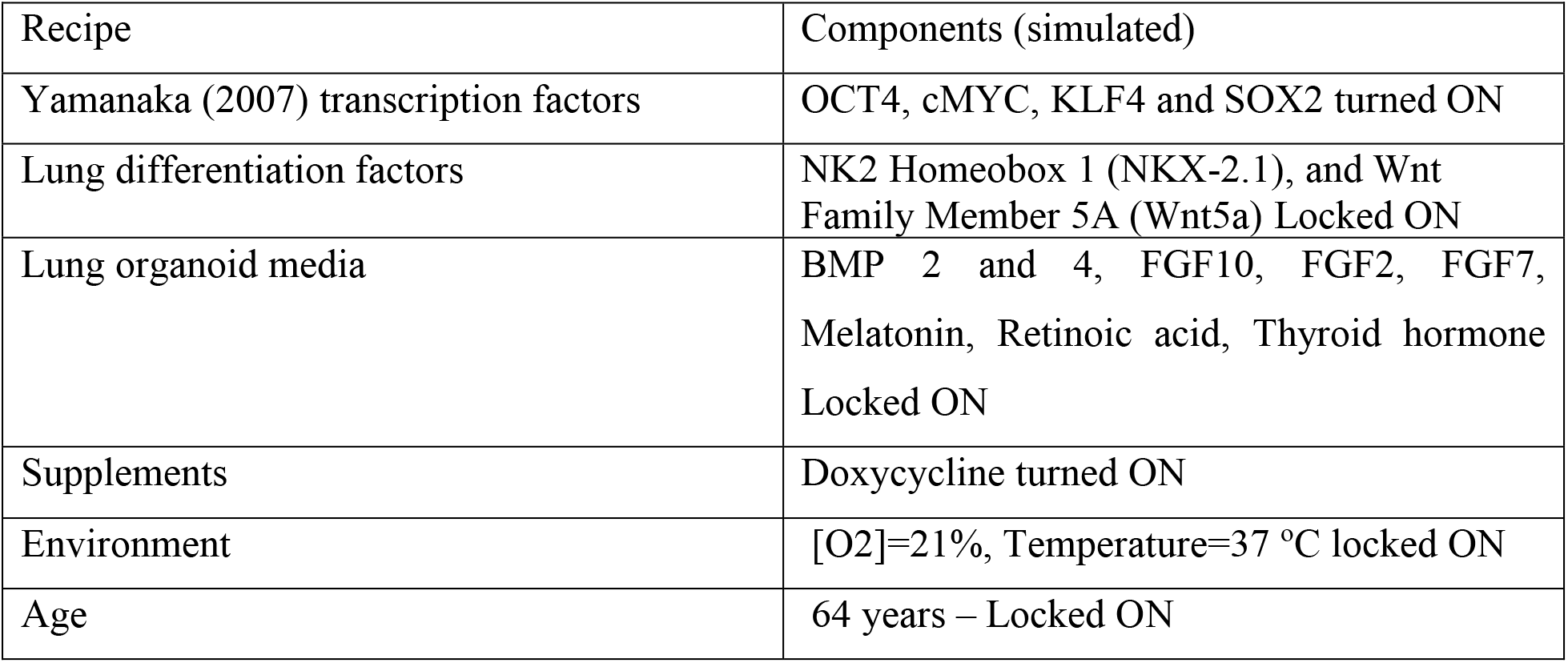
Summary of lung organoid simulation protocol

| Recipe | Components (simulated) |
| --- | --- |
| Yamanaka (2007) transcription factors | OCT4, cMYC, KLF4 and SOX2 turned ON |
| Lung differentiation factors | NK2 Homeobox 1 (NKX-2.1), and Wnt Family Member 5A (Wnt5a) Locked ON |
| Lung organoid media | BMP 2 and 4, FGF10, FGF2, FGF7, Melatonin, Retinoic acid, Thyroid hormone Locked ON |
| Supplements | Doxycycline turned ON |
| Environment | [O<sub>2</sub>]=21%, Temperature=37 °C locked ON |
| Age | 64 years – Locked ON |

### (2) Developing the adenocarcinoma of lung simulations (aiLUNG-LUAD)

Once validated, the wild type aiLUNG organoid was used to generate specific simulations of lung adenocarcinoma (aiLUNG-LUAD). To accomplish this transformation, a genetic profile of a patients’ tumor is required. Fortunately, a growing number of university laboratories and commercial companies will provide an increasingly detailed genomic analysis of an individuals’ tumor biopsy. For the purposes of this project a publicly available, HIPA compliant report containing the gene mutational profile of a biopsy with a pathological diagnosis of LUAD was obtained from a focused internet search. Data from this public domain sample report from 2018 was used as the basis for creating the aiLUNG-LUAD simulations <u>FoundationOne CDx</u> (See table 2) (Woodhouse et al., 2020). While the report does not include any demographic data other than gender, the gene profile did provide both positive and negative findings as well as a limited number (N=7) of potential therapeutic options.

**Table 2:** Biomarker/Genotypic profile obtained from a patients’ LUAD biopsy.

| Biomarker/Gene | Alteration(s) | Simulation setting(s) |
| --- | --- | --- |
| Microsatellite Instability | MSI-Stable | MSI Locked at 0 |
| Tumor Mutational Burden | TMB-Low | TMB Locked at 0 |
| ALK | EML4-ALK gene fusion<br>(GOF) | EML4-ALK Locked ON |
| TP53 | R305* mutation (LOF) | TP53 Locked OFF |
| Amplicon 11Q13 | CyclinD1, FGF19, FGF3, FGF4 | Amplicon 11Q13 Locked ON |
| CyclinD1* | Amplification | See Amplicon 11Q13 |
| FGF19* | Amplification | See Amplicon 11Q13 |
| FGF3* | Amplification | See Amplicon 11Q13 |
| FGF4* | Amplification | See Amplicon 11Q13 |
| NFKBIA | Amplification | NFKBIA turned ON |
| NKX2-1 | Amplification | NKX2-1 turned ON |

GOF=Gain of Function, LOF=Loss of Function, \*=member of Amplicon 11q13 (Schrock et al., 2019)

In the report two cancer biomarkers were evaluated. Microsatellite instability status was identified as being stable and the mutational tumor burden was classified as Low. Both biomarkers can have implications for planning optimal therapy. Genomic profiling revealed six gene amplifications and two mutations. Gene amplifications identified included CyclinD1, FGF19, FGF3, FGF4 and two mutation identified in NFKBIA and NKX2-1. A gain of function (GOF) mutation involving the ALK gene namely the EML4-ALK fusion was identified together with a loss of function (LOF) mutation in the TP53 gene (R306). In addition, several of the mutations commonly associated with LUAD in the EGFR, KRAS, BRAF, MET, RET, ERBB2 and ROS1 genes were not detected. A summary of the data obtained from the sample report is presented in Table 2 below. All experiments were carried out in triplicate using different initial conditions.

### (3) Validating the aiLUNG and aiLUNG-LUAD simulations

For the purposes of validating the simulations, two feature profiles were created. The first profile, which was used to evaluate both the aiLUNG and aiLUNG-LUAD simulations, is composed of eleven common lung cell types. These cell types including ATI and ATII precursors, Alveolar ATI and ATII cells, ATI and ATII Saccular cells plus epithelial Ciliated, Club, Goblet cells, pulmonary Neuroendocrine cells (PNEC) as well as Basal Stem Cells (BSC) were used to assess the ability of the platform to replicate both normal and cancerous LUNG organoid profiles.

A second profile containing the phenotypic and genotypic features were taken from the referenced public domain tumor expression report. These eleven features, including MSA, TMB, EML4-ALK fusion, TP53, CyclinD1, FGF19, FGF3, FGF4, NFKBIA and NKX2-1 were used to assess the ability of the platform to replicate the specific aiLUNG-LUAD profile.

### (4) Evaluating the effectiveness of single, double, and triple drug combinations on aiLUNG-LUAD

The validated aiLUNG-LUAD simulations were then used to evaluate potential drug therapies for effectiveness against the patients’ LUAD profile. The complete list of drugs including Placebo (N=43) was compiled from multiple sources including the mutation expression report <u>FoundationOne CDx,</u> (Li et al., 2020c) and FDA, Health Canada, DrugBank online, and BC Cancer Agency. A total of 43 single drugs, 903 double drug and 12,341 triple drug combinations were simulated for a total of 13,287 treatment options. A summary of the drugs evaluated are presented in Tables 3,4 and 5 below.

**Table 3:**
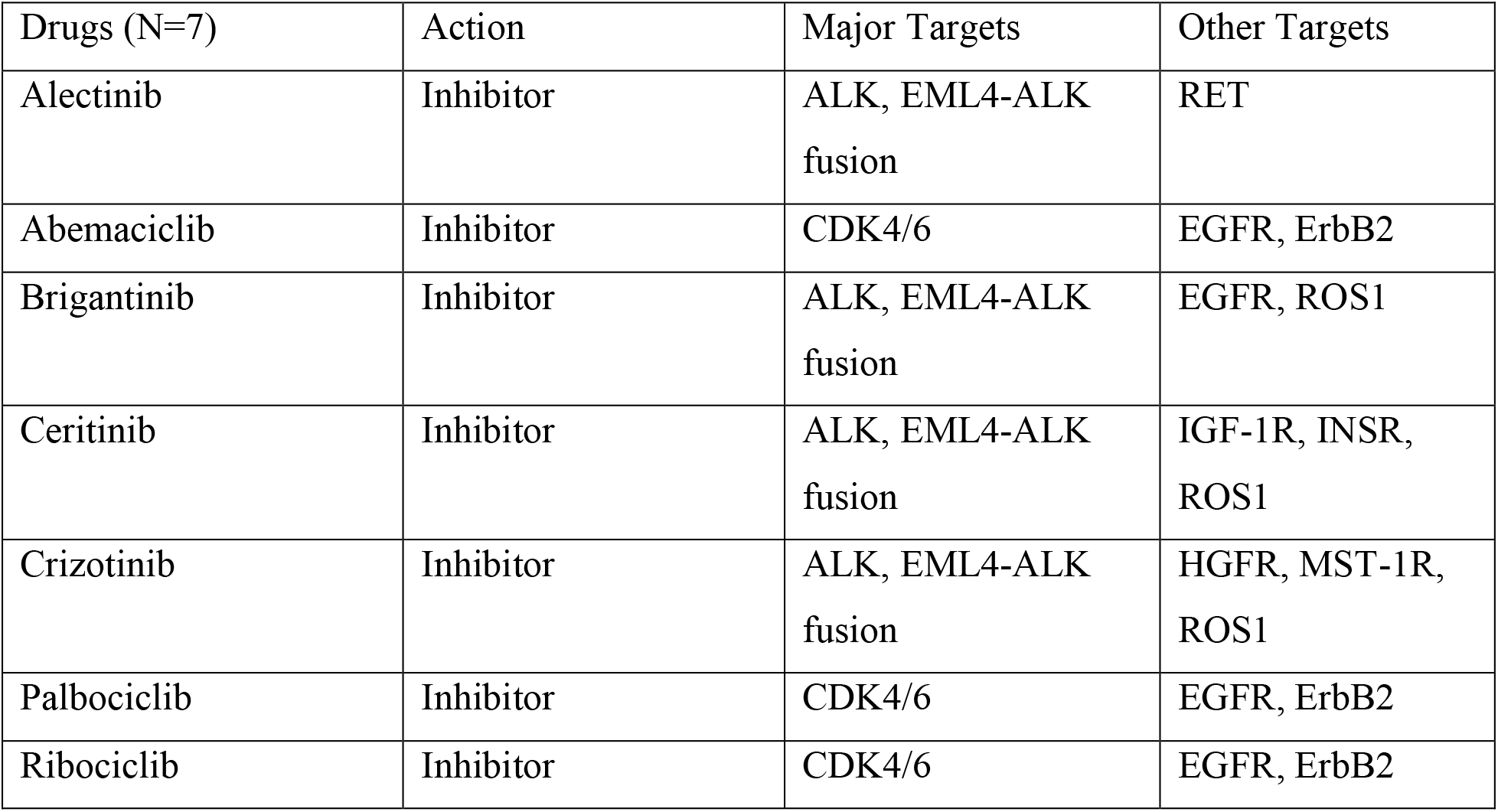
Drugs recommended by patients’ tumor profiling report

**Table 4:**
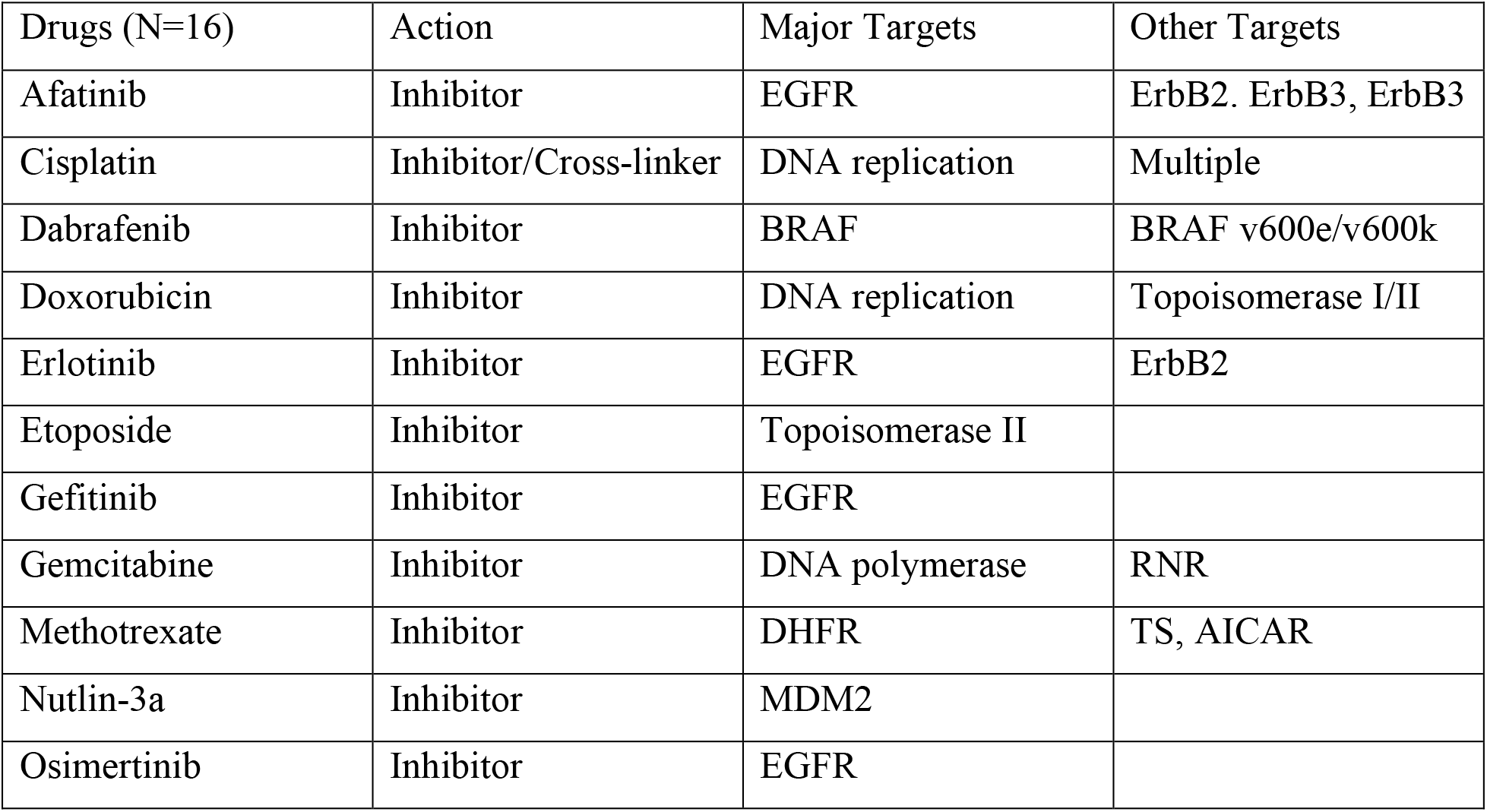
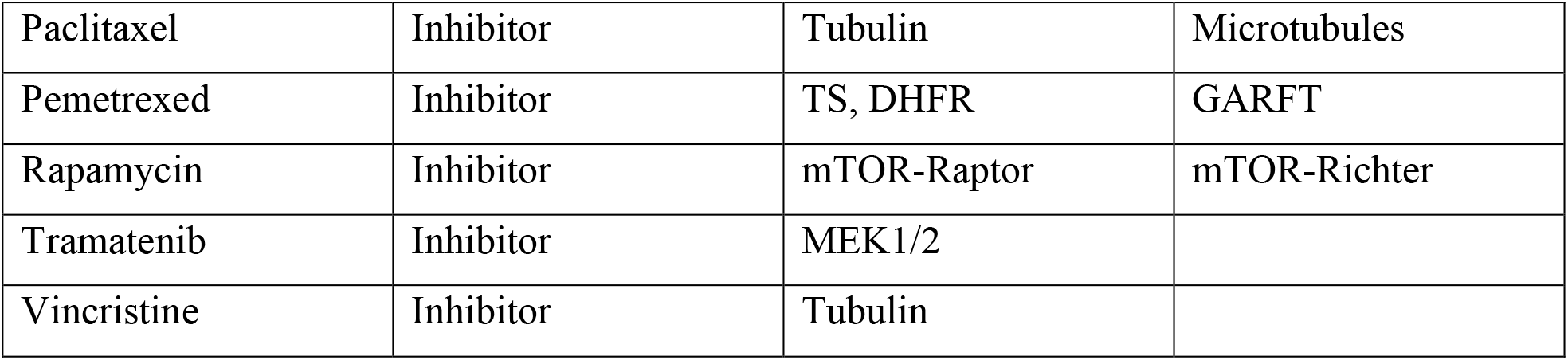
Drugs evaluated in the (Li et al., 2020c) study;

| Drugs (N=16) | Action | Major Targets | Other Targets |
| --- | --- | --- | --- |
| Afatinib | Inhibitor | EGFR | ErbB2, ErbB3, ErbB3 |
| Cisplatin | Inhibitor/Cross-linker | DNA replication | Multiple |
| Dabrafenib | Inhibitor | BRAF | BRAF v600e/v600k |
| Doxorubicin | Inhibitor | DNA replication | Topoisomerase I/II |
| Erlotinib | Inhibitor | EGFR | ErbB2 |
| Etoposide | Inhibitor | Topoisomerase II |  |
| Gefitinib | Inhibitor | EGFR |  |
| Gemcitabine | Inhibitor | DNA polymerase | RNR |
| Methotrexate | Inhibitor | DHFR | TS, AICAR |
| Nutlin-3a | Inhibitor | MDM2 |  |
| Osimertinib | Inhibitor | EGFR |  |
| Paclitaxel | Inhibitor | Tubulin | Microtubules |
| Pemetrexed | Inhibitor | TS, DHFR | GARFT |
| Rapamycin | Inhibitor | mTOR-Raptor | mTOR-Richter |
| Tramatenib | Inhibitor | MEK1/2 |  |
| Vincristine | Inhibitor | Tubulin |  |

**Table 5:**
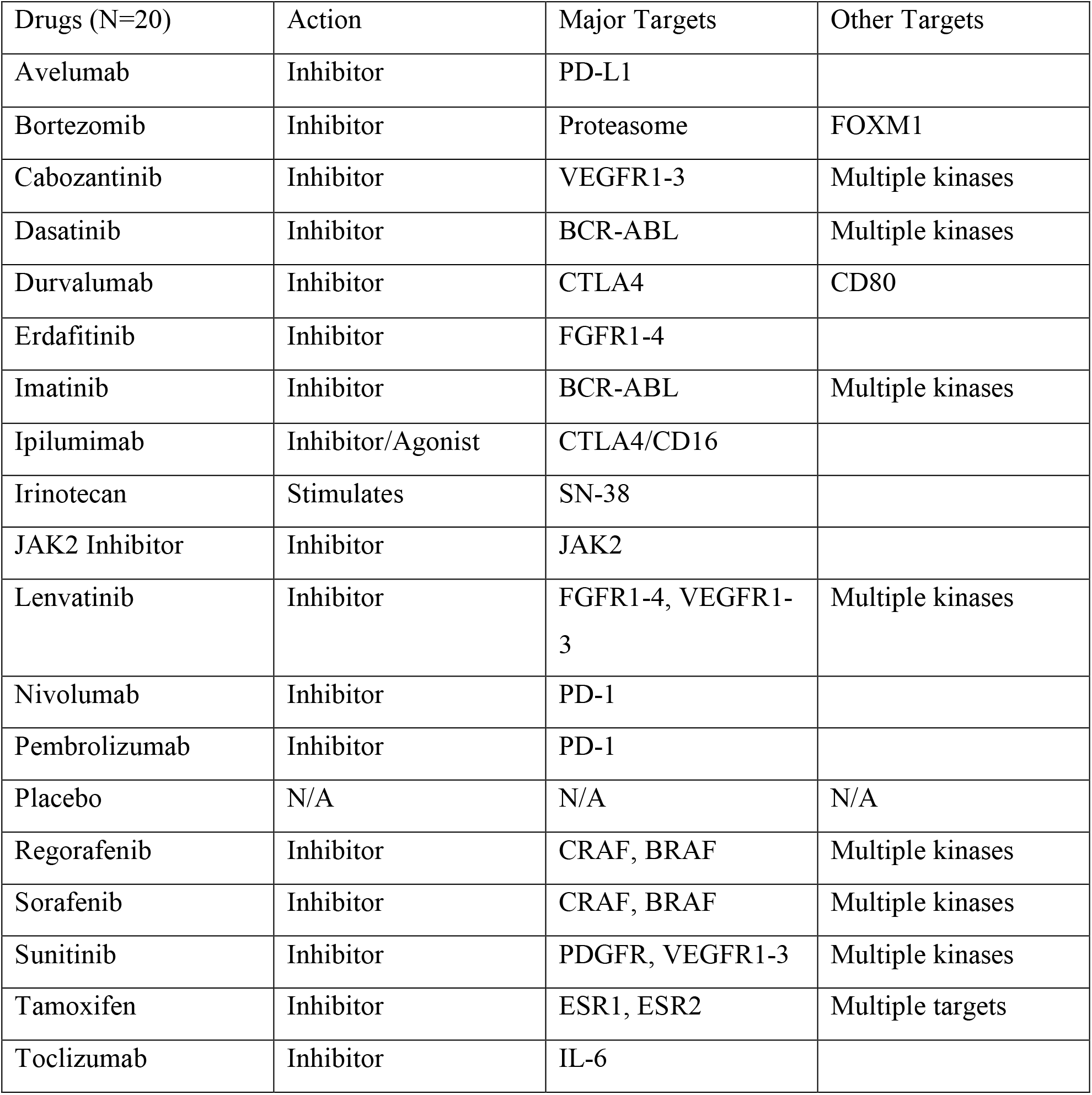
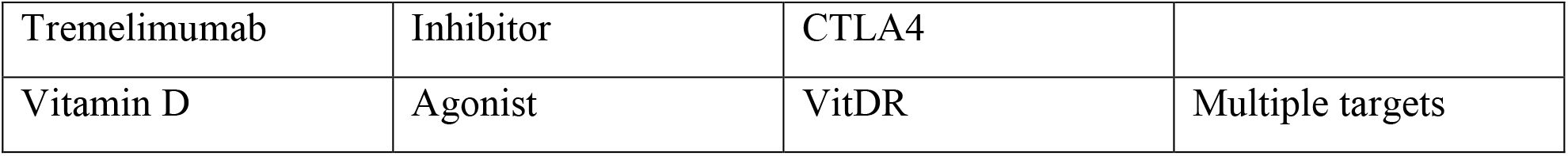
Selected targeted and multi-kinase inhibitors from multiple sources

### (5) Statistical analysis of the simulations predictions

For the aiLUNG and aiLUNG-LUAD experiments we used the unbiased binomial test to analyze simulation predictions versus the published literature. The binomial test was selected because it can compensate for prediction bias and is appropriate for determining the significance of differences between outcomes that fall into just two categories (*e*.*g*., agree vs disagree). An analysis of the updated database (6.3) identified the pre-test probability of both the positive (0.654) and negative (0.346) outcome predictions. This pretest system bias was used to adjust the binomial test prior to its use. The null hypothesis for the statistical analyses was defined as; (H0) there is no significant relationship between simulation predictions and the previously unseen peer reviewed literature. We concluded that a p value <0.05 is significant in that predicted outcomes are unlikely to have happened by chance alone. To compare between group differences (e.g., Treatment(s) vs Placebo) the two tailed paired t-test was used.

## RESULTS

### The updated DeepNEU (v6.3) database

In addition to the information contained in DeepNEU (v6.2), the updated DeepNEU database (v6.3) contains new genotypic and phenotypic concepts and relationships important for creating lung organoid simulations. For example, the previous DeepNEU database (v6.2) contained 4466 gene/proteins or phenotypic concepts and 43498 nonzero relationships while the current version (6.3) contains 4755 gene/proteins or phenotypic concepts and 42517 nonzero relationships. This represents 289 new concepts specifically relevant to developing lung organoids. Each gene/protein and phenotypic concept in DeepNEU (v6.3) has on average more than 8.9 gene/protein or phenotypic inputs and outputs. The pretest probability of a positive outcome prediction is 0.654 and the pretest probability of a negative outcome is therefore 0.346.

### The wild type aiLUNG simulations

The unsupervised aiLUNG simulations converged quickly (after 38 iterations) to a system wide steady state with no evidence of overtraining after 1000 iterations. The aiLUNG simulations successfully reproduced a wide range of lung cell types including AT I and Type II precursors, ATI and ATII cells, ATI and ATII Saccular cells plus epithelial Ciliated, Club, Goblet cells, pulmonary Neuroendocrine cells (PNEC) and lung Basal Stem cells. The probability that all 11 lung organoid cell types could be successfully reproduced by chance alone is 0.009. Importantly, these data are consistent with previous results (Li et al., 2020a) indicating that generating lung organoids from iPSC yields a diverse population of alveolar cell precursors and more mature alveolar cells. These data are summarized in Figure 1.

**Fig. 1:**
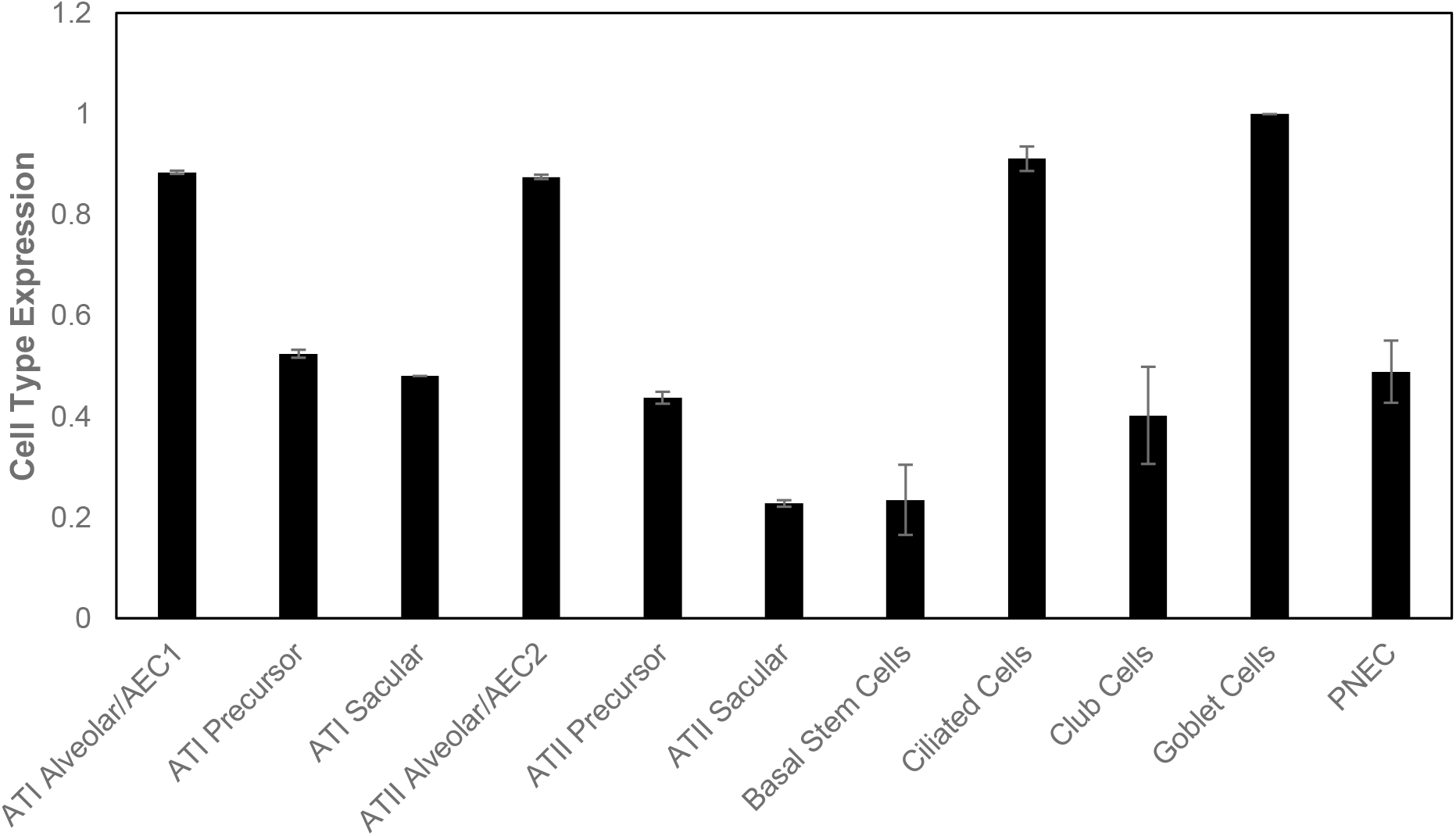
DeepNEU simulation of Lung Organoid cell types. Expression levels of aiLUNG cell types. Alveolar Type 1 cells (ATI Alveolar), Alveolar Type 2 cells (ATII Alveolar), Alveolar Type 1 precursor cells (ATI Precursor), Alveolar Type 2 precursor (ATII Precursor), Alveolar Type 1 sacular cells (ATI Sacular), Alveolar Typ2 sacular cells (ATII Sacular), Basal Stem Cells, Ciliated Cells, Club Cells, Goblet Cells and PNEC (Pulmonary Neuroendocrine Cells). The vertical axis (Y) represents the semi-quantitative Expression level of aiLUNG cell types that are estimated regarding an arbitrary base line where 0 = base line and 1= maximum expression. The horizontal axis (X) represents the individual aiLUNG cell types. Data represents mean of 3 experiments ± 99% Confidence Interval.

### The aiLUNG-LUAD lung cancer simulations

The unsupervised aiLUNG-LUAD simulations converged quickly (after 25 iterations) to a system wide steady state with no evidence of overtraining after 1000 iterations. The aiLUNG-LUAD simulations also successfully reproduced a wide range of lung cell types including ATI and ATII precursors, ATI and ATII cells, ATI and ATII Saccular cells plus epithelial Ciliated, Club, Goblet cells, pulmonary Neuroendocrine cells (PNEC) and lung Basal Stem cells. The probability that all 11 lung organoid cell types could be successfully reproduced by chance alone is 0.009. These data are summarized in Figure 2.

**Fig. 2:**
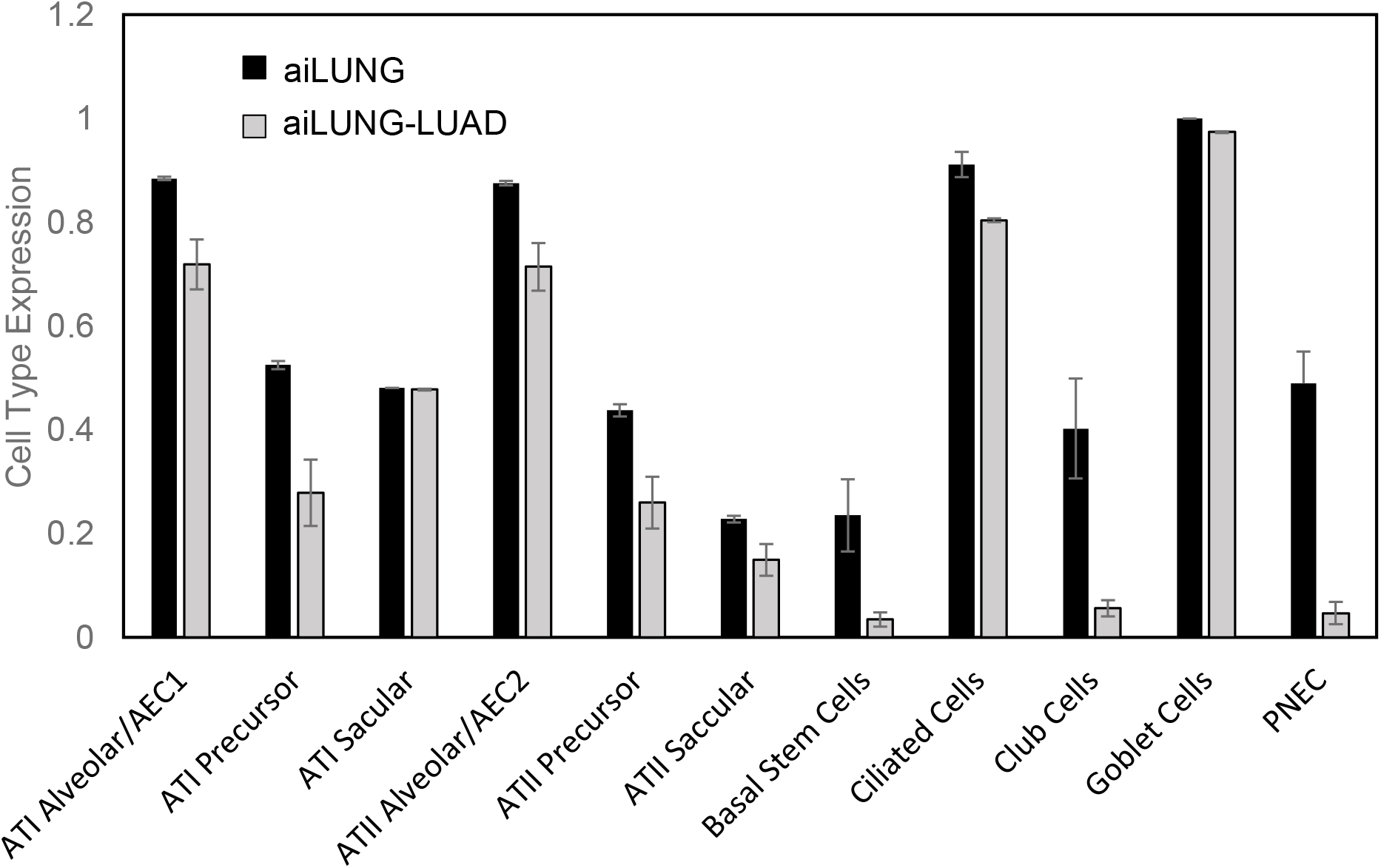
DeepNEU simulation of cell profile in aiLUNG vs aiLUNG-LUAD. Comparison of Expression of aiLUNG and aiLUNG-LUAD cell types. The vertical (Y) axis represents the semi-quantitative Expression level of cells that are estimated regarding an arbitrary base line where 0 = base line and 1= maximum expression. The horizontal (X) axis represents the semiquantitative expression level of each factor relative to arbitrary baseline. Data represents mean of 3 experiments ± 99% Confidence Interval.

A comparison of the aiLUNG and aiLUNG-LUAD simulations using the two tailed paired t-test, revealed significant differences between all lung cell types at the p<0.01 level. While all but one cell type appears to be negatively affected in the aiLUNG-LUAD simulations, the most negatively affected cells were Basal Stem Cells, Club Cells, and PNEC, all with p values <0.00001.

We also evaluated the ability of the aiLUNG-LUAD simulations to reproduce the phenotypic and genotypic features of the tumor. Consistent with the tumor profiling report, we found that there were no significant differences between the estimated MSI and TMB phenotypic features with both p values >0.5. Of the eight genotypic features, the expression of five were significantly increased relative to the wildtype lung organoid with all five p values <10E-13. These five highly significantly different genotypic factors, EML4-ALK fusion, FGF19, NKFBIA, NKX2-1 and TP53 were subsequently used in the next part of this project to evaluate the potential impact of treatments on the patients’ tumor. These data are summarized in Figure 3.

**Fig. 3:**
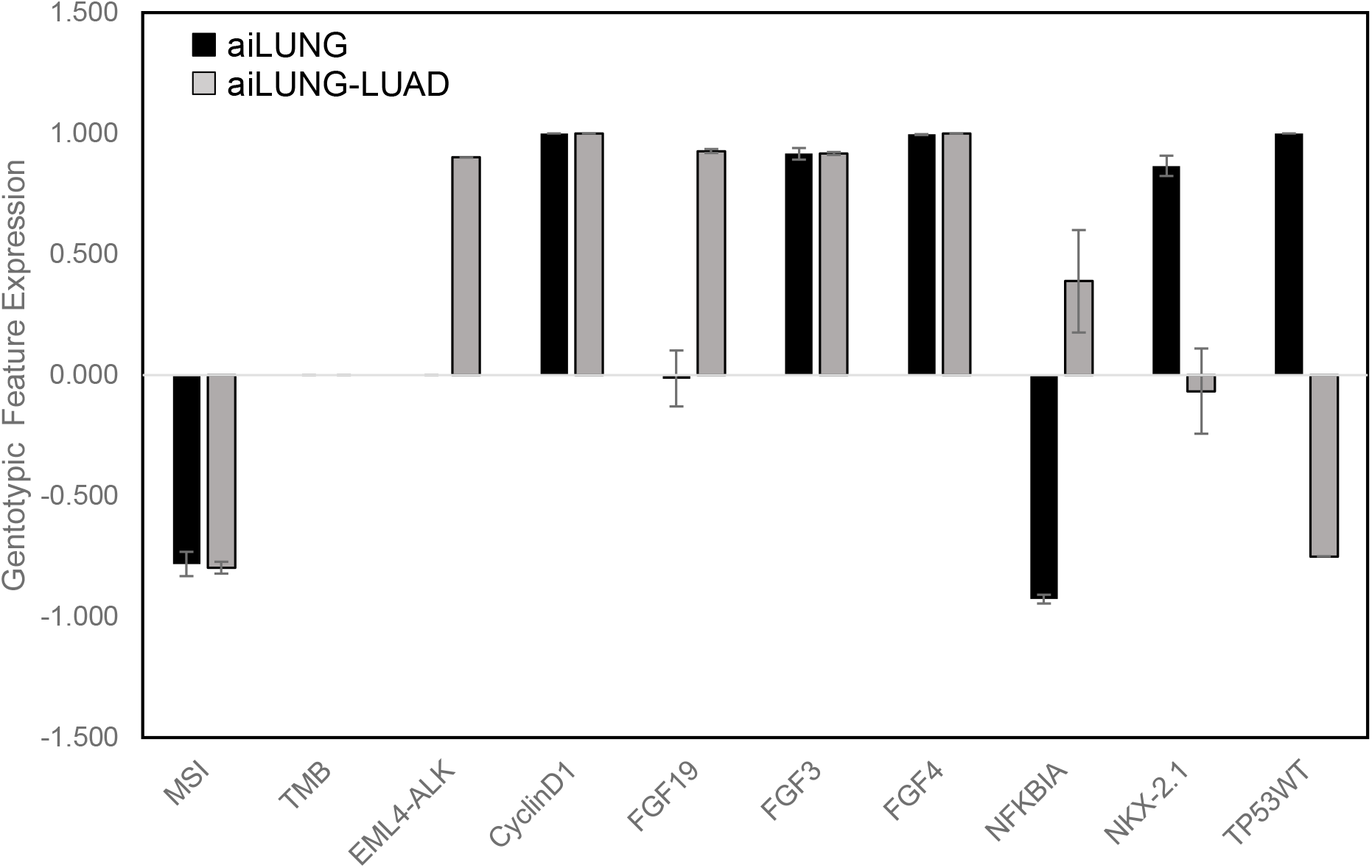
Genotypic expression profile for aiLUNG vs aiLUNG-LUAD. The vertical (Y) axis represents the semi-quantitative levels of genotypic or phenotypic factors that are estimated relative to an arbitrary base line where 0 = base line and 1= maximum expression, The X axis represents each individual mutational feature considered. Data represents mean of 3 independent experiments ± 99% Confidence Interval

### Evaluating the effectiveness of single, double, and triple drug combinations on aiLUNG-LUAD

The calculation of results for 13,287 therapeutic options took approximately 45 minutes to complete using all 8 threads of an i7Intel microprocessor addressing 32 gigabytes of RAM. All treatment predictions were compared with Placebo using the two tailed paired t-test. Using a cutoff point at p≤0.01 there were ten triple drug combinations that produced a therapeutic effect that was better than Placebo. Using a cut point at p<0.05 there were an additional 58 triple drug combinations that were different from Placebo. The number one ranked therapy overall is a combination of the IL-6 inhibitor, Tocilizumab plus the mTOR inhibitor, Rapamycin and the ALK, EML4-ALK fusion, multi-kinase inhibitor, Brigatinib with a p value of 0.003 when compared to Placebo. The highest ranking, double drug combination was Lenvatinib plus Erlotinib at #105 with a p value of 0.074 indicating it was not significantly different from Placebo. While no single drug was effective, the highest-ranking drugs were Abemaciclib, Palbociclib, and Ribociclib, all tied at #532 with p values of 0.159, once again indicating they were not significantly different from Placebo. Of interest, the mTOR inhibitor Rapamycin was a component of 9 of the top choices while Brigatinib was included in 7 of the top 10 choices. Importantly, none of the treatment options evaluated had any effect on TP53 status of the tumor.

When we evaluated the seven drugs recommended in the tumor profile report <u>FoundationOne CDx,</u> there were no single, double or triple drug combination therapies from this group that were significantly different from placebo (p>0.05). The highest-ranking options from this list were the double drug combinations of Ceritinib and Abemaciclib or Palbociclib and Ceritinib, or Ribociclib and Ceritinib, all tied for #226 with nonsignificant p values of 0.104. However, six of the seven recommended drugs were a component of at least one of the top ten options. The drug Ceritinib was not a component of the top ten options but was a component of the triple drug combination at #12. Brigatinib was the most frequent component of the top options found in 7 of the top ten. These results are summarized in Table 6.

**Table 6:**
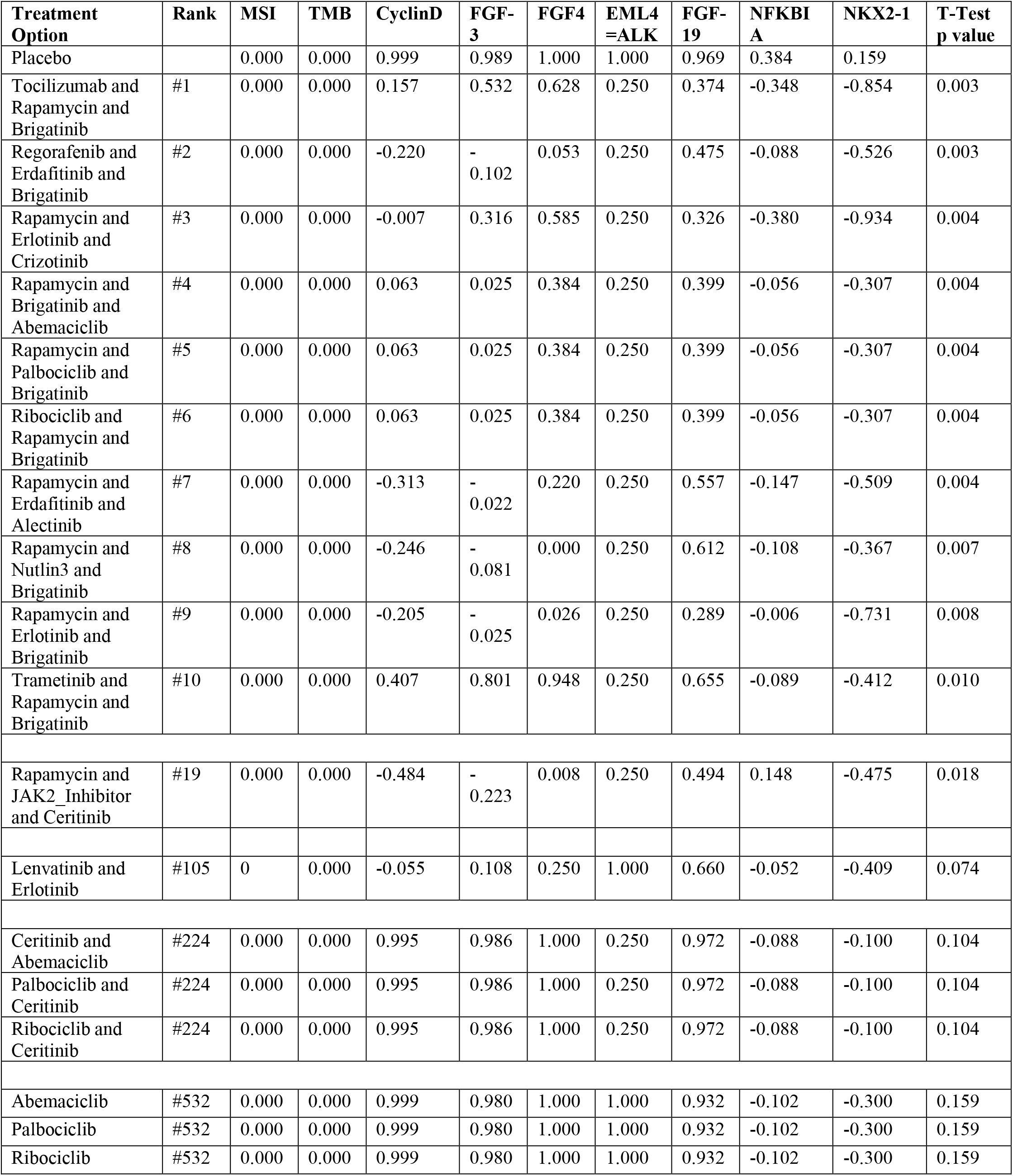
Summary of drug and drug combinations ranked by the aiLUNG-LUAD simulations

| Treatment Option | Rank | MSI | TMB | CyclinD | FGF-3 | FGF4 | EML4=ALK | FGF-19 | NFKBI A | NKX2-1 | T-Test p value |
| --- | --- | --- | --- | --- | --- | --- | --- | --- | --- | --- | --- |
| Placebo |  | 0.000 | 0.000 | 0.999 | 0.989 | 1.000 | 1.000 | 0.969 | 0.384 | 0.159 |  |
| Tocilizumab and Rapamycin and Brigatinib | #1 | 0.000 | 0.000 | 0.157 | 0.532 | 0.628 | 0.250 | 0.374 | -0.348 | -0.854 | 0.003 |
| Regorafenib and Erdafitinib and Brigatinib | #2 | 0.000 | 0.000 | -0.220 | -0.102 | 0.053 | 0.250 | 0.475 | -0.088 | -0.526 | 0.003 |
| Rapamycin and Erlotinib and Crizotinib | #3 | 0.000 | 0.000 | -0.007 | 0.316 | 0.585 | 0.250 | 0.326 | -0.380 | -0.934 | 0.004 |
| Rapamycin and Brigatinib and Abemaciclib | #4 | 0.000 | 0.000 | 0.063 | 0.025 | 0.384 | 0.250 | 0.399 | -0.056 | -0.307 | 0.004 |
| Rapamycin and Palbociclib and Brigatinib | #5 | 0.000 | 0.000 | 0.063 | 0.025 | 0.384 | 0.250 | 0.399 | -0.056 | -0.307 | 0.004 |
| Ribociclib and Rapamycin and Brigatinib | #6 | 0.000 | 0.000 | 0.063 | 0.025 | 0.384 | 0.250 | 0.399 | -0.056 | -0.307 | 0.004 |
| Rapamycin and Erdafitinib and Alectinib | #7 | 0.000 | 0.000 | -0.313 | -0.022 | 0.220 | 0.250 | 0.557 | -0.147 | -0.509 | 0.004 |
| Rapamycin and Nutlin3 and Brigatinib | #8 | 0.000 | 0.000 | -0.246 | -0.081 | 0.000 | 0.250 | 0.612 | -0.108 | -0.367 | 0.007 |
| Rapamycin and Erlotinib and Brigatinib | #9 | 0.000 | 0.000 | -0.205 | -0.025 | 0.026 | 0.250 | 0.289 | -0.006 | -0.731 | 0.008 |
| Trametinib and Rapamycin and Brigatinib | #10 | 0.000 | 0.000 | 0.407 | 0.801 | 0.948 | 0.250 | 0.655 | -0.089 | -0.412 | 0.010 |
| Rapamycin and JAK2_Inhibitor and Ceritinib | #19 | 0.000 | 0.000 | -0.484 | -0.223 | 0.008 | 0.250 | 0.494 | 0.148 | -0.475 | 0.018 |
| Lenvatinib and Erlotinib | #105 | 0 | 0.000 | -0.055 | 0.108 | 0.250 | 1.000 | 0.660 | -0.052 | -0.409 | 0.074 |
| Ceritinib and Abemaciclib | #224 | 0.000 | 0.000 | 0.995 | 0.986 | 1.000 | 0.250 | 0.972 | -0.088 | -0.100 | 0.104 |
| Palbociclib and Ceritinib | #224 | 0.000 | 0.000 | 0.995 | 0.986 | 1.000 | 0.250 | 0.972 | -0.088 | -0.100 | 0.104 |
| Ribociclib and Ceritinib | #224 | 0.000 | 0.000 | 0.995 | 0.986 | 1.000 | 0.250 | 0.972 | -0.088 | -0.100 | 0.104 |
| Abemaciclib | #532 | 0.000 | 0.000 | 0.999 | 0.980 | 1.000 | 1.000 | 0.932 | -0.102 | -0.300 | 0.159 |
| Palbociclib | #532 | 0.000 | 0.000 | 0.999 | 0.980 | 1.000 | 1.000 | 0.932 | -0.102 | -0.300 | 0.159 |
| Ribociclib | #532 | 0.000 | 0.000 | 0.999 | 0.980 | 1.000 | 1.000 | 0.932 | -0.102 | -0.300 | 0.159 |

## DISCUSSION

We have recently evaluated an earlier version of the DeepNEU machine learning platform (V5.0) for simulating the impact of SARS-CoV-2 loss of function (LOF) and gain of function (GOF) mutations in human ATI and ATII cells of simulated lung organoids (Esmail and Danter, 2021a). Previously published research also reported the ability of the DeepNEU (V5.0) platform to rapidly identify potential therapeutic targets and drug repurposing for treating COVID-19 (Esmail, 2020). The immediate predecessor of the current version V6.2, was used to successfully simulate a whole brain organoid (Esmail and Danter, 2021b). The primary objective of this project was to extend the capabilities of the evolving DeepNEU platform by evaluating the ability of the updated DeepNEU platform (V6.3) to (1) create and validate wild type lung simulations (aiLUNG-WT), (2) simulate and validate a specific gene mutational profile obtained from a patients’ tumor with a pathological diagnosis of Lung Adenocarcinoma (LUAD) and importantly to (3) evaluate single, double and triple drug combinations as potential effective and personalized therapies. As of this writing, we believe that this research represents the first successful attempt to develop and deploy simulated whole lung simulations of a human cancer mutational profile as a novel tool for precision oncology.

Consistent with previous experimental results the updated and unsupervised DeepNEU platform (V6.3) successfully reproduced literature validated profiles of aiPSC and aiLUNG genetic and phenotypic features as summarized in (Esmail and Danter, 2021a). Importantly, while V6.3 is considerably more complex than V5.0, we did not find significant differences in learning characteristics, literature validation, and performance on previously unseen data. These findings are reassuring in that they support the conclusion that there is a significant degree of internal consistency in the underlying assumptions regarding relationship information selection, data representation and the unsupervised learning approach.

The results from these experiments demonstrated that once they were literature validated, the aiLUNG-WT simulations could be modified by a combination of gene mutations and amplifications to recapitulate the commercial report regarding the patients’ tumor profile (aiLUNG-LUAD). To emphasize the importance of individual variations in individual tumors some of the more common gene mutations in LUAD including EGFR, KRAS, BRAF, MET, RET, ERBB2 and ROS1 were not detected in this patients’ tumor. On the other hand, TP53 LOF mutations are found in about 30% of LUAD while the ALK-EML4 GOF mutation is found in 7%-10% of LUAD tumors and the amplicon 11q13 (FGF3, FGF4, FGF19, CCND1 amplification) occurs in ∼13% of lung cancers. Based on this relatively uncommon tumor profile it is not surprising that standard approved therapies for LUAD may not be effective in this patient. If every cancer is different then it follows that every patient will likely need personally designed therapies to achieve the optimal results. In other words, one size does not fit all when it comes to effectively treating cancers.

Except for very few examples like Gleevec for chronic myelogenous leukemia (CML), cancer therapy often involves combinations of drugs with different mechanisms of action. The successful use of combination therapy for cancers began in 1965 when Drs Frei, Holland and Freireich introduced the POMP regimen (methotrexate, 6-mercaptopurine, vincristine and prednisone) for treating their pediatric patients with acute lymphocytic leukemia. As of this writing, the most often used two drug combinations for treating lung cancers include platins, taxols, gemcitabine, etoposide, vinorelbine, pemetrexed and irinotecan. Importantly, all these agents are traditional cytotoxic drugs with significant associated cumulative toxicity and limited success (Mokhtari et al., 2017). Less commonly three, four and even five drug combinations like FOLFIRINOX (Folinic acid + 5-FU + irinotecan + oxaliplatin) plus bevacizumab can be used to treat some advanced solid cancers resulting in some improved efficacy and barely tolerable toxicity (Chaix et al., 2014). The continued emergence of organoids and targeted, multitargeted and immune modulating agents for treating cancers will change this traditional landscape in favor of new and effective multidrug combinations (Krzyszczyk et al., 2018; Li et al., 2020b).

In the present study we used the aiLUNG-LUAD simulations to evaluate novel single, double and triple drug combinations from three broad groups. First, we evaluated a small group of seven approved targeted drugs recommended by the tumor gene profiling report for consideration by the treating physicians <u>FoundationOne CDx</u>. These seven agents inhibited either the ALK-EML4 fusion mutation or Cyclin Dependent Kinase 4/6. When this group of the seven recommended drugs was evaluated, none of the single, double, or triple drug combination therapies from this group (AA), were significantly different from placebo (p>0.05). The best options from this list of seven were the double drug combinations of Ceritinib and Abemaciclib or Palbociclib and Ceritinib, or Ribociclib and Ceritinib, all tied at position #226 in the list of 13,287 options (p=0.104, compared with placebo). However, six of the seven recommended drugs were a component of at least one of the top ten options. Brigatinib was the most frequent component of the top options found in seven of the top ten combinations, suggesting that ALK-EML4 inhibition is likely to be an important component of optimal future therapeutic options. Importantly and as expected, none of these drugs or combinations significantly impacted the TP53 LOF mutation. Going forward, this type of information could represent an important new element of the recommended therapies currently found in many commercially available genome profiling reports. A second group of potential drug candidates (N=17) that were evaluated previously in an in vitro lung adenocarcinoma (LUAD) derived organoid (Li et al., 2020c)were also evaluated in the aiLUNG-LUAD organoid simulations. This larger group is composed of more diverse traditional and targeted agents compared with <u>FoundationOne CDx</u>. In (Li et al., 2020c) these drugs were evaluated individually and combinations were not considered. Drug screening of this group in the in vitro LUAD derived organoid used the Log of the IC50 (micromolar) to evaluate anti-tumor activity over the narrow range of <10 uMol to >100 uMol drug concentrations. It is not clear that any of these drugs were active at nanomole concentrations. Consistent with these wet lab results we also found that none of the single agents from this group appeared to be effective in the aiLUNG-LUAD organoid simulations derived from the patients’ tumor profile. However, 3 drugs from this group namely Rapamycin, Erlotinib and Nutlin-3 were components of the top 10 triple drug combinations and importantly the mTOR inhibitor, Rapamycin was a component of eight of the top ten combinations. This finding indicates that mTOR inhibition is likely to be another important component of optimal future therapeutic options. Again, none of these drugs or combinations significantly impacted the TP53 LOF mutation.

The third and largest, more diverse group of drugs (N=20) is composed of targeted and multi-kinase inhibitors compiled from multiple sources. Multi-kinase inhibitors dominate in this group. Once again, none of the single drugs from this group had a significant impact on the patients’ tumor as represented by the gene profile report. However, this analysis did add the IL-6 inhibitor, Tocilizumab to the ALK-EML4 fusion inhibitor Brigatinib and the mTOR inhibitor, Rapamycin top combination. This third group of drugs also contributed the multi-kinase inhibitor, Regorafenib and the pan FGFR inhibitor Erdafitinib to Brigatinib to constitute the second rated combination. Once again, none of these drugs or combinations significantly ameliorated the negative impact of the TP53 LOF mutation.

The foregoing aiLUNG-LUAD based analysis reveals an important and unfortunate reality. Given the importance of TP53 as a tumor suppressor and the lack of more positive results from triple drug combinations that impact multiple signaling pathways but not TP53, it is unlikely that meaningful, long-term survival is possible with this specific tumor profile. The importance of TP53 in cancers is supported by the finding the TP53 mutations are found in at least 50% of all cancers and ∼30% in lung cancers (Sanz et al., 2019; Zhu et al., 2020).

One of the drugs evaluated in this project is Nutlin-3, a drug that was being developed as a potent inhibitor of MDM2, a protein responsible for inhibiting TP53 and leading to proteosome degradation. Historically, the TP53 activating effect of Nutlin like drugs (i.e. *cis*-imidazoline analogs) is most pronounced in tumors with predominantly wild type TP53. This preference for wild type TP53 likely explains the lack of effect seen with Nutlin-3 in the current experiments. A more potent Nutlin derivative RG7388 (<u>Idasanutlin</u>, ROCHE), was as of 2020 continuing in clinical development as a combination agent (Montesinos et al., 2020; Sanz et al., 2019). Another compound in Phase III clinical development is Ganetespib, a potent HSP90 inhibitor, that effects multiple signaling pathways in addition to its activity against MDM2 and CHIP that blocks TP53 deactivation and degradation (Pillai et al., 2020)).

Since its discovery in 1979, TP53 has proven to be an elusive target for drug discovery. In the last decade, several candidates have emerged that deal directly with common TP53 mutations. Two of the more advanced candidates in clinical development are APR-246 and COTI-2. Both compounds have complex mechanisms of action and at least partially act through the refolding of the mutant TP53 to a more wildtype configuration and activity (PP). Unfortunately, both drugs are still a long way from approval. A potentially viable pathway for patients with LUAD and TP53 mutations would be to enrol in future clinical trials that would allow the addition of a TP53 mutant activator to personalize combination therapy like that suggested in this paper.

## CURRENT LIMITATIONS OF AILUNG AND OTHER ORGANOID SIMULATIONS IN THE PURSUIT OF PRECISION ONCOLOGY

While there is a growing body of published data (Danter, 2019; Esmail and Danter, 2021a, b) supporting the validity of the evolving DeepNEU platform to simulate several stem cell and organoid systems, we recognize that a key limitation of this application of this emerging technology remains the degree to which the unsupervised hybrid ML platform can accurately reproduce a single patients lung adenocarcinoma using whole lung organoid simulations. These initial data reconfirm that the platform can recreate a diverse profile of previously unseen features attributed to wet lab human lung organoids in the published literature. As of this writing, simulations of a biological system like the human lung will necessarily be less complex than the actual organ. Going forward, the key to evaluating the utility of the NEUBOrg platform will be obtaining new data for learning and from ongoing validation. The data in v6.3 represents more than 21% of the human genome compared with ∼18% in the original version 5.0. Specifically, we have included many new relationships that are relevant to differentiation of stem cells into whole lung organoids. Furthermore, we have ensured through rigorous testing, that all genes reported in the commercial tumor profile report and their respective relationships are well represented. However, these advanced LUNG simulations applied to precision oncology will greatly benefit from ongoing validation including real world usage.

## FUTURE PROSPECTIVE

The immediate goal of this new DeepNEU application is to confirm these findings by expanding the simulations to as many tumor profiles as possible beyond the initial LUAD report <u>FoundationOne CDx</u>. So long as peer reviewed data exists, it should be possible to simulate virtually any tumor mutational profile from any source given the increasing capabilities of the system. A clear potential hurdle to overcome is the recommendation of unapproved drug combinations that are composed of individual drugs that have been approved. It is also possible and even likely that some combinations will be approved for indications other than the pathological diagnosis of the patient. For example, it may be that a given pathological diagnosis of LUAD is more genetically like HNSCC. Once such a finding that indicates a genotypic similarity to a different pathological type of cancer occurs, alternate treatment possibilities could be suggested.

This discussion begs the question, “who is this technology best suited for?” We want to clearly state that this application is not designed for direct patient access. The only viable route we see in 2021 is to make this information exclusively available to treating physicians or research organizations. Ideally this precision simulated cancer approach would best be made available and subsequently adopted by university/hospital tumor boards. “*A* **tumor board** *is a group of doctors and other health care providers with different specialties that meets regularly at the hospital to discuss cancer cases and share knowledge. The* **board’s** *goal is to determine the best possible cancer treatment and care plan for an individual patient*.”. Such an approach could lead to clinical trials including NofOne type trials and alternative therapies that might not otherwise have been considered. Importantly, expert tumor boards could also make these alternative therapies available to patients with appropriate and detailed informed consent.

## Data Availability

Further information and requests for resources and reagents should be directed to and will be fulfilled by the Lead Contact, Wayne R Danter

